# Short- and long-read hybrid-capture viral metagenomics reach high sensitivity and specificity with data-driven cut-offs compared with routine diagnostics in CSF and BAL specimens

**DOI:** 10.64898/2026.10.27.26366582

**Authors:** Mustafa Karatas, Lize Cuypers, Lies Laenen, Mandy Bloemen, Annabel Rector, Justine Maes, Marthe Vanrenterghem, Marc Van Ranst, Elke Wollants, Stefanie Desmet, Jelle Matthijnssens

## Abstract

**Background:** Hybrid-capture viral metagenomics improves sensitivity over untargeted metagenomics, but its performance across sequencing platforms and the definition of data-driven analysis cut- offs using real clinical samples have not been systematically evaluated. This study benchmarks the hybrid-capture workflow on mock and clinical specimens across short- and long-read platforms while introducing a new, reproducible approach for setting pathogen-calling cut- offs.

**Methods:** The Twist Comprehensive Viral Research Panel was used with both AVITI short-read and Oxford Nanopore Technologies (ONT) long-read sequencing on 12 mock cerebrospinal fluid (CSF) external quality assessment (EQA) panel samples and on 14 CSF and 21 bronchoalveolar lavage (BAL) clinical specimens. Data-driven cut-offs for pathogen calling were derived using Latin hypercube sampling across genome coverage, abundance, and bleed-ratio parameters, then validated on the independent EQA datasets.

**Results:** After cut-off application, both platforms detected 22 of 23 expected viral targets in EQA samples (95.7% detection-level sensitivity) with 100% specificity in negative controls. In clinical specimens, AVITI corroborated 26 of 30 (86.7%) and ONT 25 of 30 (83.3%) routine viral detections while maintaining 100% diagnostic specificity. Sensitivity was 100% for CSF but lower in BAL, particularly for low viral load targets, and all detections below a Cq value of 32 were identified by both platforms. Metagenomics also identified a potentially clinically relevant pathogen not tested by routine testing in two CSF samples.

**Conclusion:** Hybrid-capture viral metagenomics performed comparably on AVITI and ONT for routine viral targets, with AVITI showing slightly higher genome recovery. In addition, the newly developed data-driven cut-off selection strategy offers a practical, reproducible method other laboratories can apply to their own workflows.

## Introduction

Conventional diagnostic testing for viral infections relies mainly on targeted molecular assays and syndromic panels. These methods are sensitive for the pathogens they cover, but additional testing may be required when the initial diagnostic hypothesis is incorrect or when the causative virus is not included in the assay. Viral metagenomic next-generation sequencing (mNGS) provides a broader alternative and has identified clinically relevant viral infections missed initially by conventional testing in cerebrospinal fluid (CSF) specimens.^1–4^ Metagenomic sequencing of bronchoalveolar lavage (BAL) specimens has also supported pathogen detection in severe pneumonia, although most studies have focused primarily on bacterial and fungal infections. ^5,6^ A remaining limitation of viral mNGS is reduced sensitivity for low-abundance targets, particularly in specimens containing large amounts of host nucleic acids.^7^

Hybrid-capture, also referred to as target enrichment, increases the proportion of viral reads by enriching viral nucleic acids before sequencing. This can improve read recovery and sensitivity, particularly when viral nucleic acids are present at low abundance within a high human background. ^8^ Mourik *et al*. showed sensitivity and specificity of ≥95% with limits of detection around 50–500 copies/mL across synthetic controls, reference virome mixtures and a limited set of clinical samples (n=8).^8^ While it should be noted that hybrid-capture panels are inherently limited to their predefined target space, the 120-mer probe design of the Comprehensive Viral Research Panel (Twist Bioscience, USA) tolerates up to 20% sequence divergence reportedly, allowing detection of novel members of known virus families through conserved genomic regions.^9^

Beyond enrichment, the choice of sequencing platform may affect read recovery, genome coverage, turnaround time and laboratory implementation. Hybrid-captured libraries can be analyzed using either short-read or long-read sequencing. ^10^ Beyond the platform choice, result interpretation and cut-off selections remain the main hurdles. This is further complicated by low read counts that may arise from index hopping, barcode misassignment, or cross-contamination during the wet-lab handling of the sample. Consequently, cut-offs are highly dependent on the laboratory, sequencing technology, enrichment approach, and sample type. While some data-driven approaches for background filtering and pathogen calling exist,^11,12^ most studies define and apply their cut-offs for virus presence rather arbitrarily without data-driven approaches.

Therefore, few studies have evaluated hybrid-capture viral metagenomics on the same specimens using both short- and long-read sequencing, and cut-off selection is rarely described using reproducible approaches. In addition, BAL specimens remain comparatively understudied in viral metagenomics, while CSF has received more attention. We designed a two-phase study in which hybrid-capture viral metagenomics was first evaluated using external quality assessment panels and was then applied to 14 CSF and 21 BAL clinical specimens. Routine microbiological results obtained during standard clinical care were used as the reference standard for routine-target performance analyses. Using the clinical dataset, we developed a Latin hypercube sampling-based approach to derive platform-specific cut-offs based on genome coverage, abundance, and bleed-ratio parameters. These cut-offs were then applied to the blinded EQA datasets and to the clinical cohort to compare AVITI short-read and ONT long-read sequencing with conventional routine and reflex PCR results.

## Methods

### **1.** Study Design

Our study comprises two complementary components: a controlled analytical evaluation using external quality assessment (EQA) panels and a retrospective clinical evaluation using 14 cerebrospinal fluid (CSF) and 21 bronchoalveolar lavage (BAL) specimens from patients with suspected central nervous system infections or severe pneumonia. AVITI short-read and Oxford Nanopore Technologies (ONT) long-read sequencing were performed in parallel for all samples. Platform-specific pathogen-calling cut-offs were derived from the clinical CSF and BAL dataset and subsequently applied without modification to the EQA datasets for independent analytical evaluation. Metagenomic findings in the clinical specimens were compared with routine diagnostic results and, where available, reflex PCR results (**Figure 1**).

**Figure 1.**
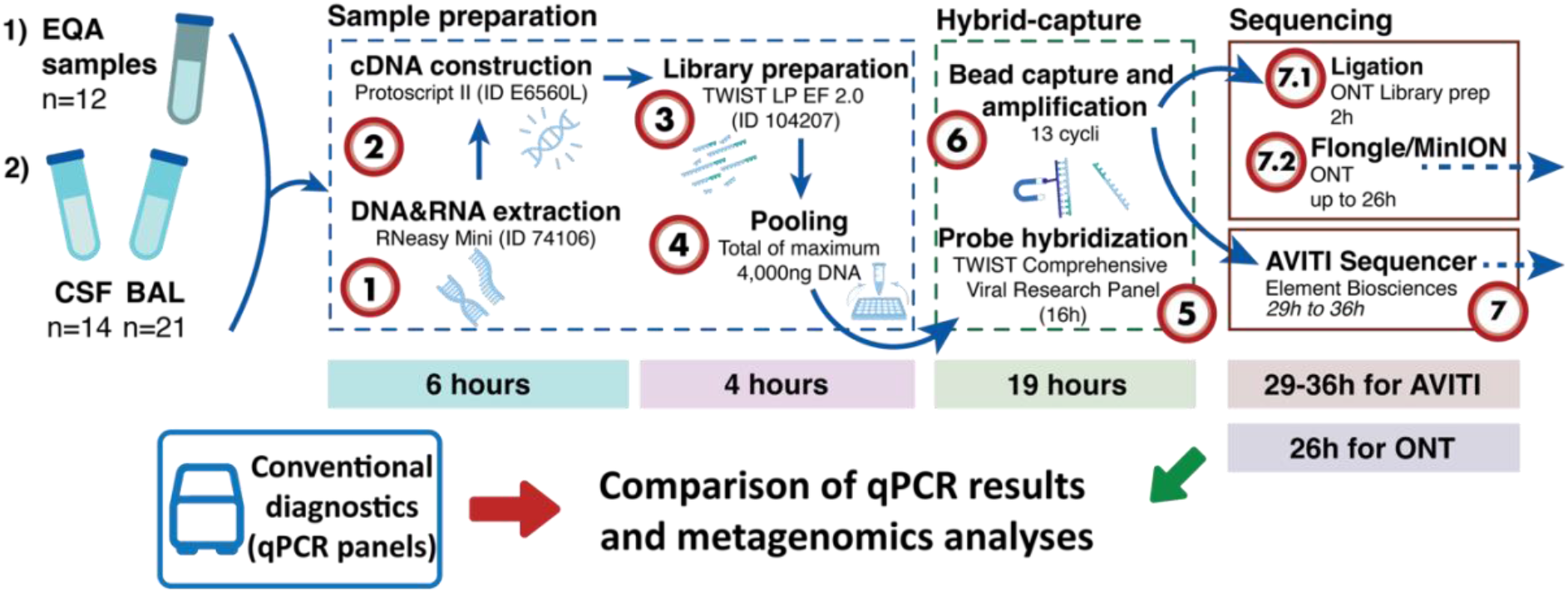
Schematic overview of the study, including sample processing, sequencing, and downstream analysis for AVITI and ONT. The timeline shows platform-specific average turnaround times.

### **2.** EQA panel analyses

To evaluate the workflow under controlled conditions with predefined viral load, we participated in the 2024 and 2025 viral metagenomics EQA programmes organized by Quality Control for Molecular Diagnostics (QCMD, UK). Each annual panel comprised six lyophilized or transport-medium-based mock CSF specimens containing defined viral targets or negative controls. After sequencing, virus-assigned read counts, genome coverage, and sequencing depth were extracted for all expected sample-virus pairs. Virus assignments not expected in the corresponding sample were also recorded before application of the study-specific operational cut-offs and classified according to the QCMD reference composition. The complete distribution of expected and non-expected viral signals is provided in **Supplementary Table 1**. The platform-specific cut-offs derived from the clinical dataset were subsequently applied without modification to the EQA datasets, which were not used for parameter optimization.

### **3.** Sample selection for CSF and BAL study

Clinical specimens were selected retrospectively from residual material stored at -20°C at the clinical laboratory of UZ Leuven. The CSF sample collection comprised 14 specimens collected in 2023 and 2024 from patients with suspected central nervous system infection, including seven samples with a viral diagnosis by routine testing, two with a bacterial detection, three with no causative agent identified by routine testing and no increased cell counts on cytology, and two additional cases selected after consultation with infectious disease specialists because an infectious origin remained clinically suspected. The BAL sample collection comprised 21 specimens from patients hospitalized with severe pneumonia in 2024 or 2025, including 14 samples in which at least one viral pathogen was identified by routine respiratory testing and seven samples in which no viral pathogen was detected. This cohort was designed as a retrospective virus enriched clinical evaluation cohort rather than a consecutive diagnostic cohort.

### **4.** Routine and reflex testing for BAL and CSF

Routine molecular diagnostic results were used as the reference standard for routine-target performance analyses. Among the seven CSF samples with a routine viral diagnosis, five had been tested using the QIAstat-Dx Meningitis/Encephalitis panel (QIAGEN)^13^, whereas two had been tested using in-house pathogen-specific PCR for EBV in accordance with the University Hospitals Leuven accredited in-house laboratory protocol.^14^ All BAL samples were analyzed using an in-house broad respiratory panel targeting 29 viral, bacterial and fungal pathogens after NucliSens easyMAG extraction.^15^ Additional metagenomic findings (and false negatives) were re-evaluated by PCR tests when residual sample material was available, which is referred to as “reflex PCR” throughout the text. For CSF, reflex PCR was performed for EBV or HHV-6 in two samples. For BAL, reflex PCR was performed for EBV, HHV-6 and HHV-7. EBV and HHV-6 were tested using in-house qPCRs, whereas HHV-7 was tested using a targeted endpoint PCR covering 12 amplicons with in-house accredited laboratory protocol of UZ Leuven.^14^ Four initially positive detections by routine testing, *Enterovirus* in sample B1, HSV-1 and RSV in sample B3 and HSV-1 in B16 were found to be negative by metagenomics and reflex testing, and were therefore excluded from the sensitivity and specificity calculations as well as cut-off selections and discussed separately since this may be due to sample degradation between routine and metagenomic tests conducted. All samples and PCR tests as well as Cq values were given in ***Supplementary methods Table 1***.

**Table 1.**
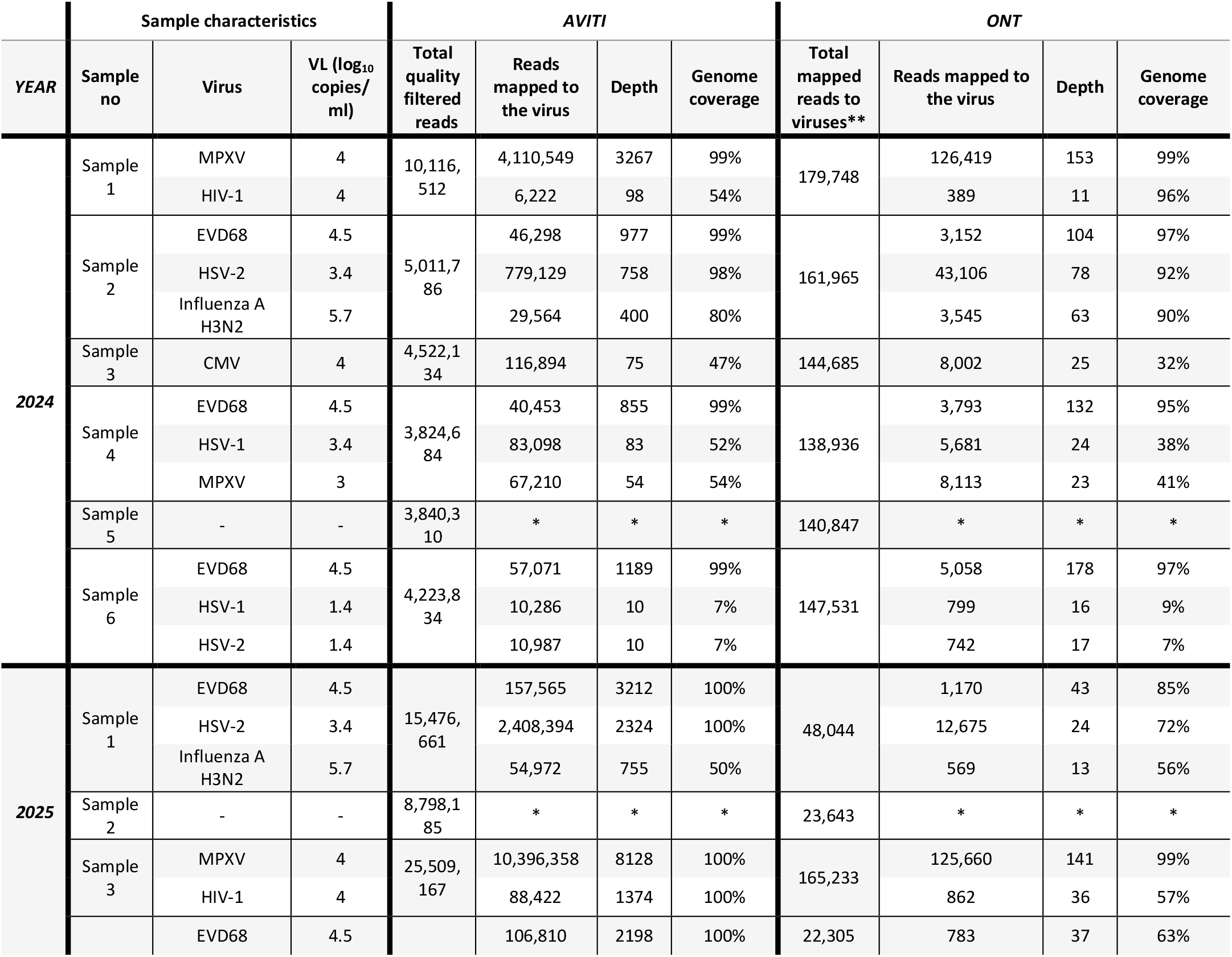
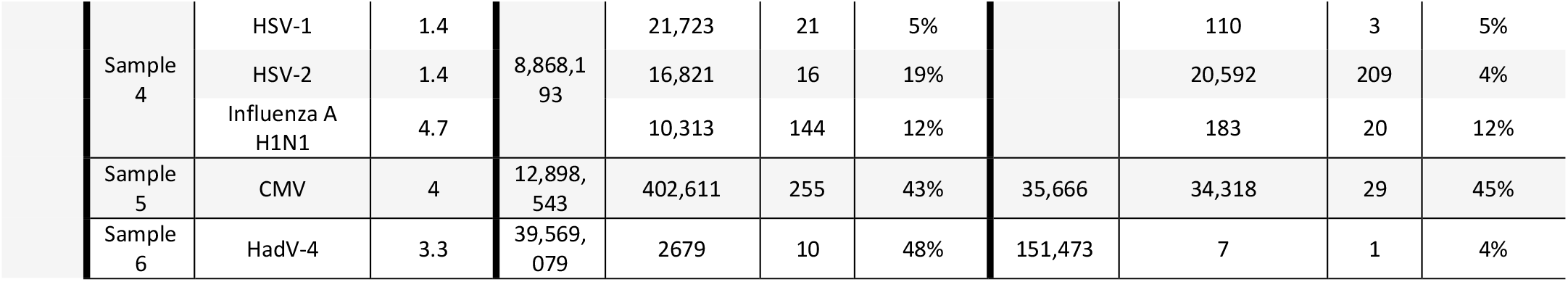
Results for expected viral detections in EQA samples after hybrid-capture per sequencing platform. VL: viral load as communicated by QCMD. *\*Multiple virus reads were assigned to range of viruses in negative samples, which is explained in the text and shared in Supplementary* table 1*. **Genome Detective performs an initial assembly followed by a remapping step; here we report the final mapped read counts to viruses in the sample*.

### **5.** Metagenomics analyses

Wet-lab processing was performed as previously described.^16^ Briefly, total nucleic acids were co-extracted from CSF and BAL specimens using the RNeasy kit (QIAGEN, 74106), followed by cDNA synthesis with ProtoScript II (New England Biolabs, E6560L) and library preparation with Twist Library Preparation EF Kit 2.0 (Twist Bioscience, USA, catalog number 104207). Libraries were enriched using the Twist Comprehensive Viral Research Panel (Twist Bioscience, USA). ^9^ After 16 hours of hybridization and 13 cycles of post-capture PCR, enriched CSF and BAL library pools were sequenced separately on an AVITI sequencer (Element Biosciences, USA) and on R10 MinION/GridION flow cells using a Mk1C and GridION device. Bioinformatic detection and quantification of viral pathogens for AVITI was performed using EsViritu v0.2.3^17^ with no internal cut-offs. For ONT sequencing, enriched libraries were prepared using the previously explained ligation-based Nanopore workflow^18^, basecalled and demultiplexed in MinKNOW using the super-accurate model, and filtered at Q-score ≥10 before analysis with *Genome*

*Detective Virus Tool* (version 2.21.4). ^19^ Several viral taxa were excluded from pathogen-calling optimization and clinical performance analyses before application of the operational cut-offs. These included signals attributable to high-titer non-study libraries sequenced in the same run, such as sapovirus and yellow fever virus; known laboratory or reagent-associated contaminants, such as circovirus-like genomes and parvovirus NIH-CQV; probable taxonomic misassignments involving closely related viruses or endogenous viral elements, including several animal herpesviruses and retroviruses; and viruses outside the predefined clinical scope of acute central nervous system (CNS) infection and lower respiratory tract infections, such as anelloviruses and papillomaviruses. These signals were retained in the unfiltered output and are reported, together with the reason for exclusion, in **Supplementary Table 2**.

### **6.** Cut-off selection process for metagenomics

Platform-specific cut-offs were derived from the clinical CSF and BAL dataset using Latin hypercube sampling in R. Each sample-virus pair was treated as a separate detection event. For each platform, 100,000 candidate cut-off sets were generated, each defined by three thresholds: 1) minimum genome coverage, 2) minimum abundance, and 3) minimum bleed ratio, used to filter low-level cross-sample signals associated with high-abundance taxa. Abundance was expressed as reads per million quality-filtered reads for AVITI and as raw virus- assigned read counts for ONT. Candidate cut-off sets that retained detections in negative clinical samples were excluded, and the remaining sets were ranked according to predefined criteria first prioritizing diagnostic specificity followed by detection sensitivity (explained in detail in ***Supplementary Methods***). The 100 highest-ranked sets were carried forward to assess the consistency of parameter selection and performance, and the best-performing set was selected for the main analysis. The selected set and the other 99 highest-ranked sets were applied without modification to the EQA datasets, which were not used for cut-off optimization. The selected operational cut-off set was also used for the final analysis of the clinical CSF and BAL samples.

“Detection sensitivity” was defined as the proportion of expected sample-virus pairs recovered by metagenomics taking routine clinical diagnostics (and reflex testing) detections as the reference. “Sample sensitivity” was the proportion of positive samples in which all expected viruses were recovered. “Diagnostic specificity” was the proportion of negative samples with no metagenomic detection after filtering. “Off-target removal” was the proportion of non-expected sample-virus signals removed after cut-off application. The parameter ranges, ranking criteria and cut-off selection procedure are described in detail in the ***Supplementary Methods***.

## Results

### 1. Expected EQA targets generated reads, but non-expected signals were widespread

We first assessed raw viral read recovery using two EQA panels comprising 12 EQA CSF samples, including negative controls. Across both panels, 23/23 (100%) of the expected sample-virus targets were present at concentrations ranging from 1.4 to 5.7 log_10_ copies/mL. Both AVITI and ONT recovered reads assigned to all expected targets, although read counts, sequencing depth and genome coverage varied substantially across viruses and concentrations (**Table 1**).

Interpretation was complicated by reads assigned to non-expected viruses in both positive samples and negative controls. These signals frequently corresponded to viruses present at high abundance in another sample processed within the same sequencing run, including MPXV, HSV-2, and were consistent with potential cross-sample bleed, index hopping or low-level background contamination. The complete distribution of expected and non-expected viral signals is provided in **Supplementary Table 1**.

Among the 23 expected sample–virus pairs, excluding the two negative EQA samples, we examined whether genome coverage and normalized read abundance reflected the input viral load. Viral load showed a significant positive Spearman correlation with genome coverage for both AVITI (ρ = 0.451, p = 0.031) and ONT (ρ = 0.555, p = 0.006). In contrast, viral load was not significantly correlated with normalized read abundance for either AVITI (ρ = 0.104, p = 0.636) or ONT (ρ = −0.016, p = 0.942) (Figure 2).

**Figure 2.**
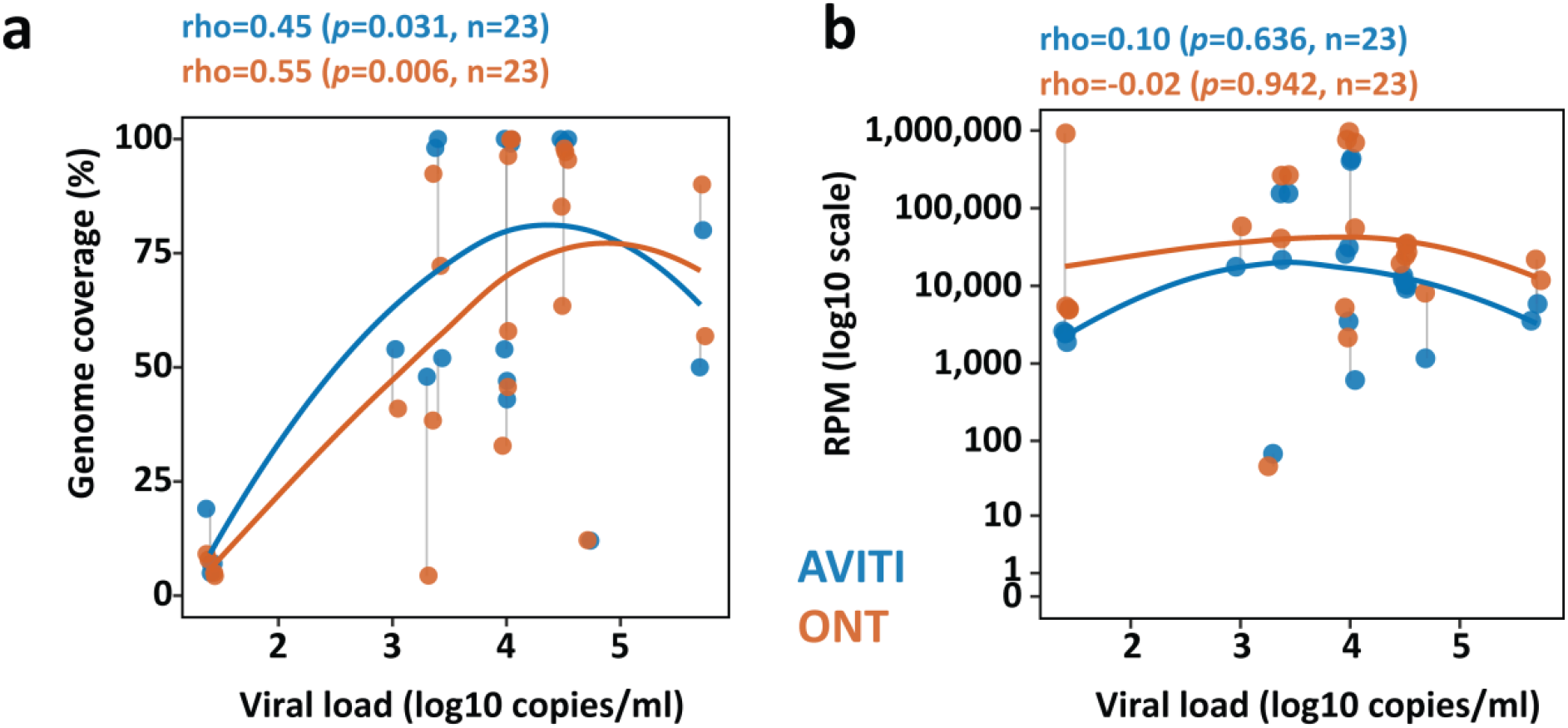
**Comparison of viral load (as determined by QCMD) with metagenomics parameters from the EQA samples**. **a.** Comparison of genome coverage (%) and viral load of each detection. **b.** Reads per million filtered reads (RPM) per sequencing method compared with viral load, normalized read abundance was calculated using platform-specific denominators: total quality-filtered reads for AVITI and total mapped viral reads for ONT. Both figures only include the expected 23 detections, while negative samples were excluded from these analyses.

Thus, although hybrid-capture sequencing recovered reads from all expected targets, read presence was not sufficient to define a positive result or exclude false-positive detections, and genome coverage was a better correlating metric with viral load. We therefore aimed to develop platform-specific cut-offs incorporating abundance, genome coverage and the relative signal of each virus across samples to control for high viral load sample bleeding and/or cross-contamination.

### 2. Platform-specific cut-offs removed most non-expected signals in EQA dataset

As we show in the previous section, probe-based viral metagenomic sequencing can detect very low-level signals, but not every signal should be interpreted as “presence” of a virus. Low read counts may arise from wet-lab contamination, index hopping, cross-sample contamination, or non-specific mapping. Therefore, we aimed to select stringent, data-driven cut-offs to distinguish reproducible viral detections from other false signals based on results of the routine diagnostics as reference, while preserving sensitivity for true positives. To define these cut-offs, we sampled 100,000 possible quasi-random parameter combinations separately for AVITI and ONT. Each combination sampled three cut-off values: I) how much of the viral genome was covered, II) how abundant the viral reads were (as read count or RPM), and III) whether the signal was strong enough relative to the highest signal for the same virus within the sample type, referred to as the bleed ratio. These candidate cut-off sets were evaluated on the clinical CSF and BAL datasets, and the best-performing parameter sets of 100 combinations were selected (*see methods*). These 100 best-performing parameters sets were then applied to the EQA datasets for external validation (Figure 3a). The selected optimal cut- offs were intentionally stringent (see methods and *Supplementary methods*) but remained permissive enough to retain high analytical sensitivity. The top 100 ranked parameter sets for each platform fell in a relatively narrow range for each parameter compared to the sampling range, which may indicate that the ranges are potentially a high-performing solution ranges rather than random combinations performing well as outliers (Figure 3b-c).

**Figure 3.**
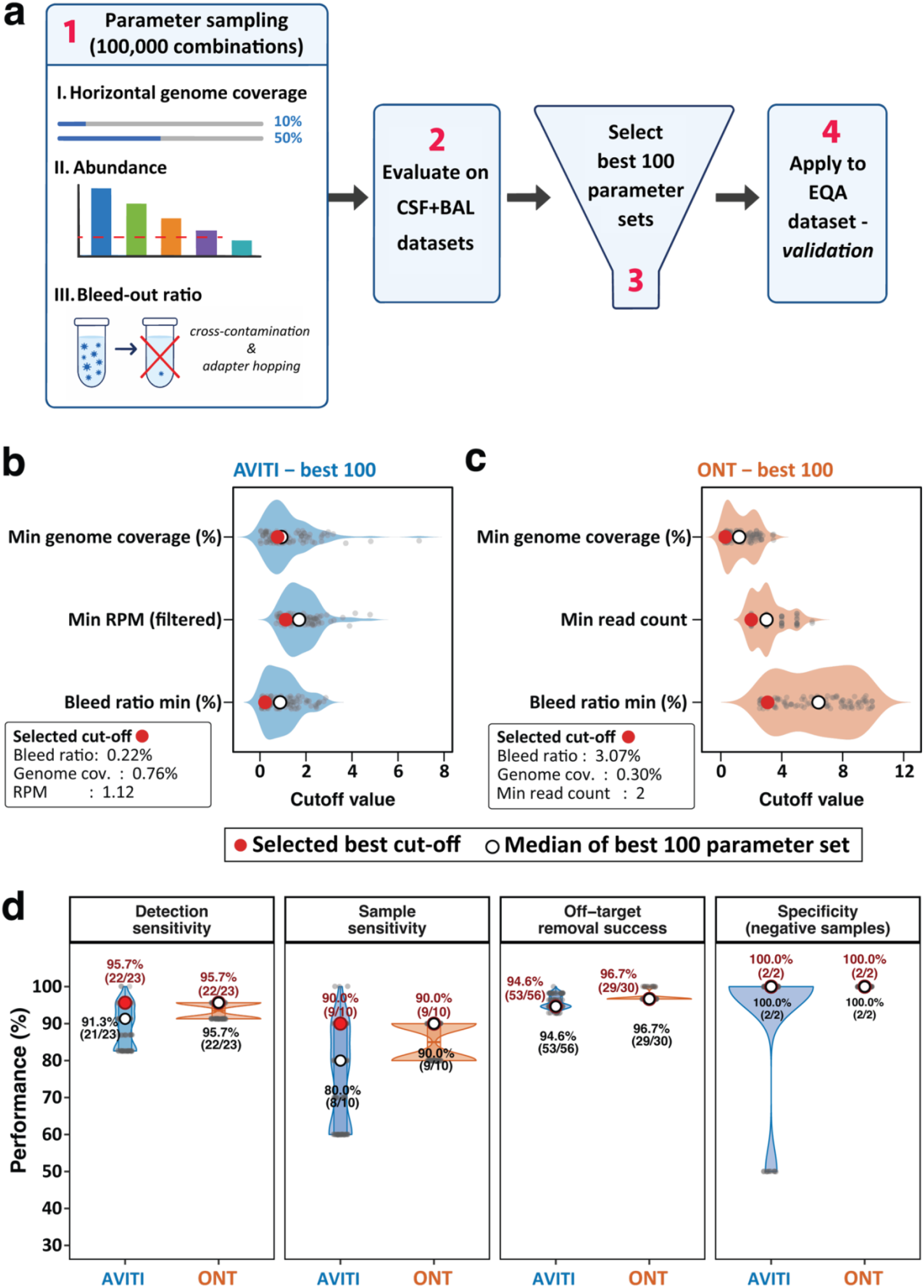
a. Schematic overview of cut-off selection and validation with the EQA dataset. **b-c.** Top 100 internally ranked cut-off sets for AVITI (b) and ONT (c). Grey points show individual parameter sets, white open circles show the top-100 median parameter values, and red circles show the selected operational cut-offs. **d.** Detection sensitivity, sample sensitivity, observed off-target removal, and negative-sample specificity in the 12 EQA samples across the top 100 cut-off sets. White circles show performance of the median cut-off set; red circles show performance of the selected operational cut-off set. “Detection sensitivity”: detection capability of metagenomics when all virus detections are pooled regardless of the samples that they were identified from. “Sample sensitivity”: Detection is deemed successful when every virus in one sample is detected. “Off-target removal” was defined as the proportion of observed raw non-expected sample–virus signals removed after cut-off application. “Diagnostic specificity”: Metagenomics identifying negative samples as negative.

After application of the selected operational cut-offs on the EQA dataset, 22 of the 23 expected viral targets remained positive on each platform. AVITI did not retain human adenovirus 4 (HadV-4) in 2025 Sample 6, whereas ONT did not retain HSV-2 in 2024 Sample 6. The AVITI HadV-4 detection comprised 2,679 reads and 48.0% genome coverage but failed the bleed-ratio criterion because a high-abundance adenovirus-positive clinical sample was co- sequenced in the same run and therefore provided the denominator for this calculation. Post- hoc sequence-level comparison supported the HadV-4 signal as a genuine detection distinct from the potential donor sample, indicating that it was removed by a conservative bleed-ratio filter. Among non-expected sample–virus combinations with an observed raw sequencing signal, the selected cut-offs removed 53 of 56 AVITI signals (94.6%) and 29 of 30 ONT signals (96.7%). Both platforms correctly classified the two negative EQA samples after filtering, corresponding to 100% sample-level specificity (2/2).

### 3. Sensitivity is sample type and viral load dependent

Because operational pathogen-calling cut-offs should reflect clinical sample matrices rather than artificial control material alone, the cut-offs were derived from the clinical CSF and BAL dataset. Using these cut-off values, we assessed routine-target performance in the same cohort of 14 CSF and 21 BAL specimens, using routine molecular diagnostic results as the reference standard. Detection-level sensitivity was defined per individual pathogen detection, whereas sample-level sensitivity required recovery of all expected viruses within a given sample.

Overall, AVITI corroborated 26 of 30 routine pathogen detections (86.7%), while ONT corroborated 25 of 30 (83.3%) (**Figure 4a**). Both platforms detected all routinely targeted pathogens in CSF samples (7/7). In BAL, detection-level sensitivity was 82.6% for AVITI (19/23) and 78.3% for ONT (18/23), while sample-level sensitivity was 71.4% for AVITI (10/14) and 64.3% for ONT (9/14). No routine-negative clinical samples showed retained viral detections after cut-off application, corresponding to 100% diagnostic specificity.

**Figure 4.**
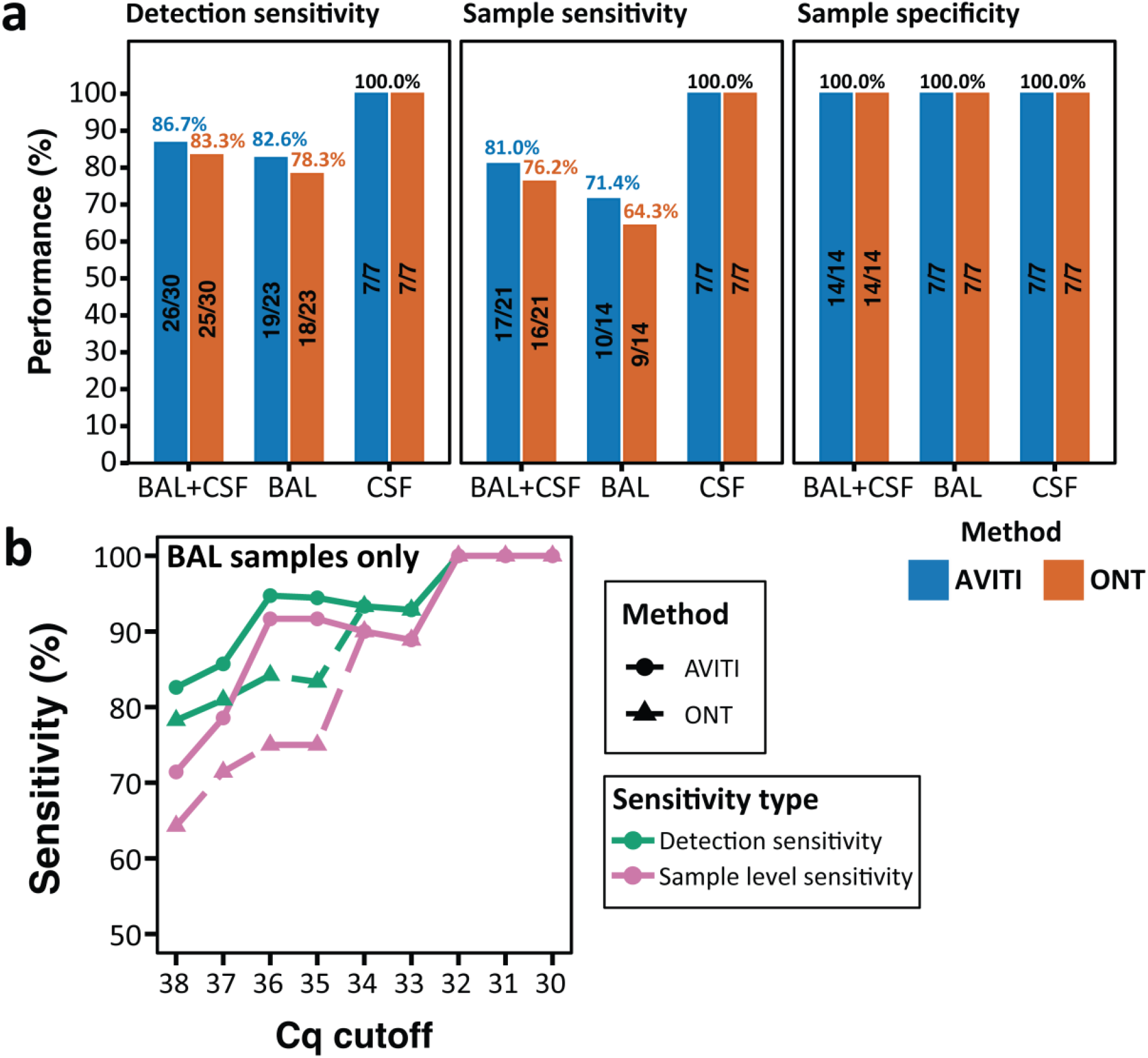
Clinical performance of cut-off-filtered viral metagenomics for pathogens tested in routine diagnostics. **a.** Detection sensitivity, sample sensitivity, and sample specificity after applying the selected operational cut-offs to clinical CSF and BAL samples. Results are shown for all samples combined, BAL only, and CSF only. Labels indicate percentage and numerator/denominator. **b.** Sensitivity in BAL samples according to routine PCR Cq threshold. Detection sensitivity reflects individual sample-virus targets; sample sensitivity reflects the percentage of samples in which all expected targets were detected.

Because sensitivity was lower in BAL samples, we further examined the missed routine detections. All missed detections had Cq values above 32, with 3 of 4 AVITI misses and 2 of 5 ONT misses occurring above Cq 36. Below a Cq threshold of 36, AVITI reached 90% detection sensitivity; below a Cq threshold of 34, both platforms reached more than 90% detection sensitivity (**Figure 4b**). These findings indicate that clinical sensitivity was influenced by both viral load and sample type.

### 4. Viral metagenomics can identify overlooked pathogens and remove the biases of stepwise testing

To further examine discordant and additional viral findings, we summarized all PCR-tested sample-detected virus pairs together with the corresponding cut-off-filtered AVITI and ONT metagenomic results (Figure 5a). Among routinely targeted viruses, most detections were recovered by both sequencing platforms: 24 of 30 routine PCR-positive sample-virus pairs were detected by both AVITI and ONT. AVITI detected 26 of 30 routine targets overall, whereas ONT detected 25 of 30. Four routine PCR-positive detections were negative on reflex testing and were therefore shown separately (dark red in Figure 5a); these discordant reflex-negative results may reflect loss of detectable signal after sample handling or freeze-thawing, rather than true failure of metagenomic detection.

**Figure 5.**
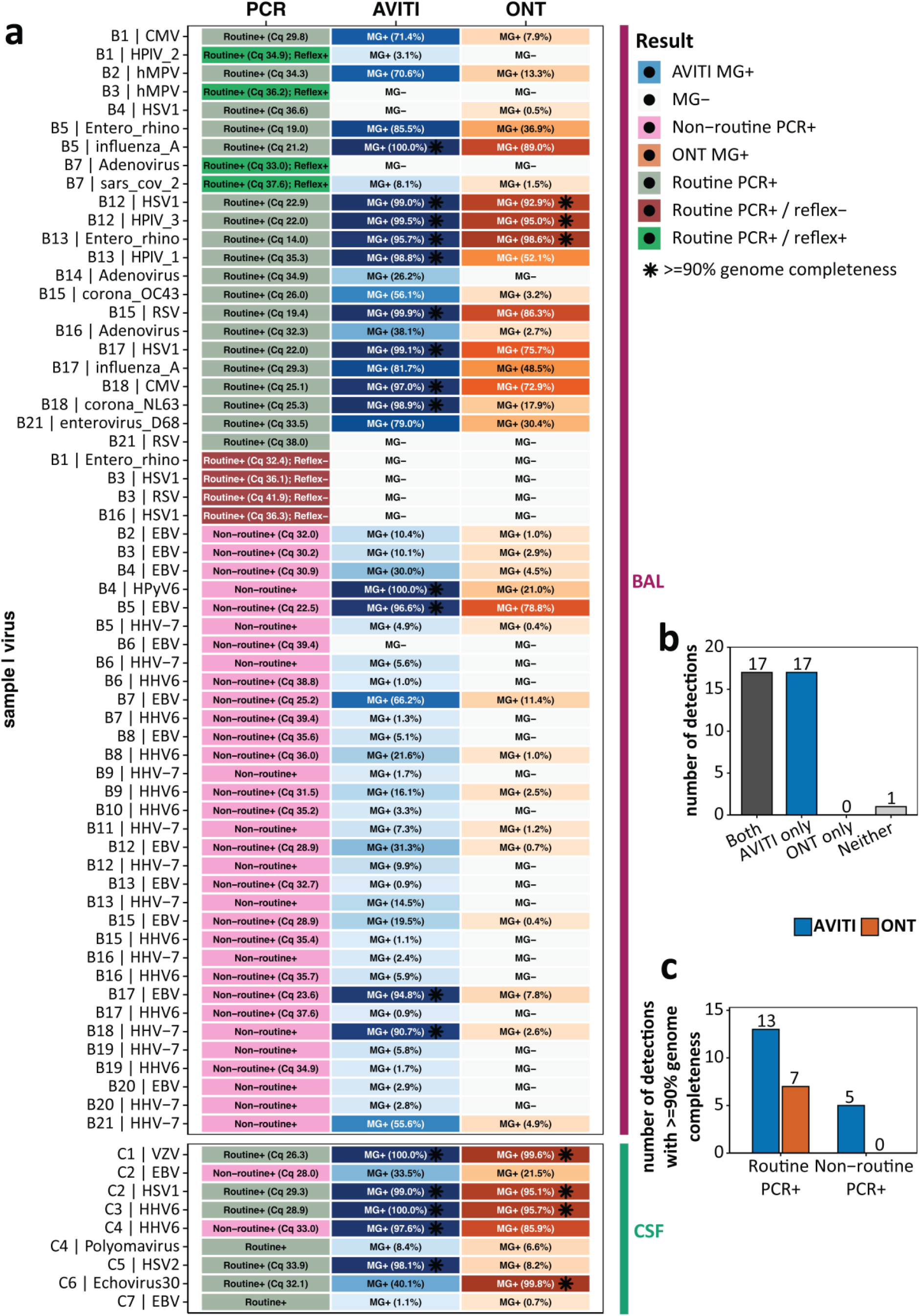
Routine diagnostics (PCR) and cut-off-filtered metagenomics concordance and genome recovery by sequencing platform. Heatmap showing PCR-based sample-virus detections in BAL and CSF specimens, with corresponding cut-off filtered metagenomic results for AVITI and ONT. Each row represents one sample-virus pair; columns show PCR result, AVITI result, and ONT result. PCR labels indicate whether the target was identified by routine testing, routine testing with reflex result, or non-routine PCR, with Cq shown where available. Metagenomic labels show whether the virus was detected after application of the selected platform-specific cut-offs, with genome completeness shown in parentheses. Black asterisks mark detections with >=90% genome completeness. Blue indicates AVITI-positive metagenomic detections, orange indicates ONT-positive detections, and gray indicates no metagenomic detection after filtering. **b.** Number of non-routine PCR-positive detections recovered by both platforms, by AVITI only, by ONT only, or by neither platform. **c.** The number of viral detections yielding near-complete genomes (>=90% genome completeness) for AVITI and ONT, stratified by routine and non-routine PCR viral targets.

Metagenomic findings also prompted additional reflex PCR testing (shown as non-routine or reflex in Figure 5), particularly for highly prevalent herpesviruses in BAL samples, including EBV, HHV-6 and HHV-7. Among 35 non-routine PCR-positive detections, 17 were recovered by both sequencing platforms, while an additional 17 were recovered by AVITI only; no non-routine detection was recovered by ONT alone, and one was missed by both methods. Thus, AVITI showed slightly higher routine-target sensitivity and recovered substantially more additional low-abundance non-routine detections than ONT (Figure 5b). Genome recovery also differed between platforms. Across the dataset, AVITI recovered 18 near-complete genomes (>=90% genome completeness), compared with 7 by ONT (Figure 5c). Among routine PCR-positive detections, near-complete genomes were recovered for 13 AVITI detections and 7 ONT detections. Among non-routine PCR-positive detections, AVITI recovered 5 near-complete genomes, whereas ONT recovered none.

The clinical relevance of additional EBV, HHV-6 and HHV-7 detections in BAL samples remains uncertain. In contrast, two CSF samples illustrated the potential value of broader viral detection (C2 and C4). In sample C4, routine testing identified JC polyomavirus with 2.87 log_10_ copies/mL. Metagenomics recovered JC polyomavirus at low genome completeness (8.4%) and additionally detected HHV-6 with a near-complete genome, which was confirmed by reflex PCR (Figure 5a). These findings suggest that stepwise CSF diagnostic algorithms may miss CNS-relevant viral targets that are detectable by broad metagenomic testing.

## Discussion

In this study, we benchmarked hybrid-capture viral metagenomics using both short-read (AVITI) and long-read (ONT) sequencing across mock EQA samples and clinical CSF and BAL specimens, and developed a data-driven approach to define cut-offs for both platforms. We achieved 95.7% (22/23) detection-level sensitivity on EQA samples, 100% (7/7) sensitivity on clinical CSF routine targets, and high overall clinical routine-target sensitivity, reaching 86.7% for AVITI and 83.3% for ONT. For CSF, the observed sensitivity was higher than the pooled sensitivity reported in a recent meta-analysis of mNGS for infectious meningoencephalitis,^20^ suggesting hybrid-capture enrichment may improve sensitivity in CSF samples, supporting the literature work on mock samples.^21^ Sensitivity in BAL samples was lower, reaching 82.6% for AVITI and 78.3% for ONT at individual virus detection level, which we speculate that higher host DNA background in this sample type may be reducing capture efficiency during hybridization. An additional reason may be potential viral signal loss from freeze-thaw cycles prior to processing, which we show that four lower viral load detections were not detected when reflex-testing was conducted.

Despite expanding clinical use of viral mNGS, pathogen calling still relies on ad-hoc thresholds such as minimum read counts (often 1–2 reads), 2–3 non-overlapping regions of coverage, fixed RPM or percentage of total reads cut-offs, or simple fold-change over negative controls, with little consensus or formal justification. ^22,23^ These heterogeneous and permissive rules may contribute to high sensitivity but widely variable specificity, which can drop to about 26% in severe pneumonia cohorts and shows broad inter-study spread in diagnostic test accuracy meta-analyses.^24,25^ Contamination-focused work shows that cross-sample bleed, index hopping and reagent “kitome” DNA are pervasive in viral mNGS, yet most studies still address them qualitatively rather than embedding them into quantitative decision thresholds. In our study, we present a Latin hypercube sampling approach that jointly optimizes genome coverage, abundance and bleed-ratio parameters to select best-performing cut-off values and potentially a plausible operating range, enabling more balanced comparisons between sequencing platforms instead of ad-hoc cut-offs. These cut-offs are not intended to be universal but are laboratory- and workflow-specific; when wet-lab protocols or bioinformatic pipelines change, repeating the same optimization procedure can help redefine appropriate operational thresholds in a transparent, data-driven way.

Multiple CSF mNGS studies have shown that unbiased sequencing can reveal unexpected viral etiologies missed by stepwise testing, including Parvovirus B19, enteroviruses, hepatitis E virus and even vesiviruses in patients initially labelled as having idiopathic or autoimmune CNS disease.^1,26^ In our cohort, one sample similarly illustrated this added value: metagenomics identified HHV-6 with higher genome completeness alongside JC polyomavirus with lower genome completeness (which is the only virus reported in the routine diagnostics results), with HHV-6 only confirmed by reflex PCR. In BAL, we frequently observed EBV, HHV-6 and HHV-7, echoing mNGS studies that detect lower-respiratory herpesviruses in roughly one-third of immunocompetent pneumonia patients and associate them with longer hospital stays and potentially with immune status of the patients.^27^

To our knowledge, this is the first direct head-to-head comparison of short-read and long-read sequencing performed on the same hybrid-captured viral libraries across both mock EQA panels and clinical specimens. Prior studies comparing sequencing technologies have either combined hybrid capture exclusively with short-read platforms or applied ONT to untargeted workflows, where sensitivity at low viral loads consistently lags behind Illumina-based approaches.^21^ In our study, the performance gap was narrower than the literature suggests; both platforms achieved similar routine-target sensitivity and 100% specificity, though AVITI showed an advantage in low-abundance non-routine detection and genome recovery. This is potentially due to extra steps involved before ONT sequencing as well as sequencing depth difference between technologies rather than the sequencing itself. On the other hand, ONT’s commonly cited speed benefit is also substantially narrowed in hybrid-capture workflows, where the 16-hour hybridization step dominates turnaround regardless of sequencer choice. Intrinsic differences in sequencing depth between the short-read sequencing run (6.9M reads per sample in our study) and the long-read flow cell (20k reads per sample in our study) should nonetheless be kept in mind when comparing results.

With sensitivity and specificity now more systematically addressable, cost and turnaround time remain the practical challenges for routine clinical implementation of viral metagenomics. Newer short-read platforms with lower read output but shorter runtime, advances in hybridization chemistry and emerging PCR-free on-flow-cell hybrid-capture approaches are collectively pushing total workflow time below two days.^28,29^ On the other hand, long-read technologies can be analyzed in real time which can provide data access before completion of the sequencing run. Additionally, short-read technology used in this study requires batching of multiple samples, whereas long-read technologies may use less number of samples being pooled together. On the other hand, even if sequencing may be conducted on a low-throughput sequencing platform for a low number of samples, hybridization still requires batching of multiple samples. Overall, while both platforms show technical feasibility, the choice between them should be informed by the tradeoffs, costs, and batching needs when multiple samples have to be sequenced.

Our study has limitations that should be acknowledged. The clinical cohort is small, and samples were processed from frozen material, which may have contributed to nucleic acid degradation and reduced sensitivity for some targets which we show with reflex PCR testing. Additionally, sample selection was based on the availability of residual material, diagnostic testing on CSF samples was not standardized, as tests were performed according to the clinician’s request as part of standard clinical care. Furthermore, cut-offs were derived and evaluated using the same clinical cohort, which may have produced optimistic performance estimates. However, their performance on independent EQA panels suggests that the cut-offs retained discriminatory ability beyond the derivation cohort. That said, missed detections occurring at Cq values above 32 and only in BAL samples suggest that our sensitivity estimates reflect genuinely difficult samples rather than workflow failure, making them conservative rather than optimistic benchmarks for prospective clinical use. Moreover, our study offers a reproducible cut-off selection approach usable to any laboratory running similar workflows. Additionally, to our knowledge, this is the first parallel short-read and long-read comparison on the same hybrid-captured libraries across both controlled EQA samples and real clinical CSF and BAL samples.

## Data Availability

Human reads filtered raw datasets can be found in Sequence Read Archive (SRA) in the Bioproject number PRJNA1475316. Scripts to reproduce the analyses and figures can be found on https://github.com/Matthijnssenslab/clinicalVMGS_UZL_KUL.

## Acknowledgements

We thank Sarah Gillemot and Graciela Andrei for their help on HHV-7 testing, and Kurt Beuselinck and laboratory technicians of Laboratory Medicine UZ Leuven for conducting the reflex-testing for the relevant samples.

## Ethics

The study protocol was accepted by the Ethics Committee Research UZ/KU Leuven (Belgium) under reference number S72724.

## Use of artificial intelligence tools

Codex 5.6 Sol and GPT-6 Astra were used to assist with proofreading, paraphrasing, data analysis and visualization, and assessing the manuscript for consistency. All AI-assisted outputs were reviewed and verified by the authors and edited where necessary. The authors retain full responsibility for the analyses, interpretation and final manuscript.

