## Supplementary methods for "Short- and long-read hybrid-capture viral metagenomics reach high sensitivity and specificity with data-driven cut-offs compared with routine diagnostics in CSF and BAL specimens"

### 1.1. Clinical specimen selection criteria and routine diagnostic and reflex PCR details

Clinical specimens were selected from residual diagnostic material available after completion of routine testing. Inclusion required sufficient residual volume for nucleic acid extraction and availability of corresponding routine diagnostic results. Specimens were prioritized to evaluate analytical and clinical performance across routine-positive, routine-negative, low-abundance, and co-detection scenarios. For BAL specimens, selection additionally prioritized samples reflecting the complexity of respiratory material, including mixed viral detections and samples with potentially competing high- and low-abundance viral signals.

Reflex PCR assays were performed only for selected additional metagenomic findings when residual material was available. Assay details, including target region, primer/probe references, amplification format, and positivity criteria, are provided in Supplementary Table Z.

For performance analyses, routine molecular diagnostic results were used only to define expected routine viral targets. Reflex PCR results were used to evaluate additional metagenomic findings and were not used to redefine the original routine diagnostic status of the specimen.

### 1.2. Cut-off identification via sampling

#### 1.2.1. Parameter sampling

Operational cut-offs were optimized separately for AVITI and ONT using Latin hypercube sampling implemented in R with the *lhs* package. Each row represented one viral target in one clinical sample. Sample–virus pairs were classified as expected when the virus was reported by routine diagnostic testing and as non-expected when the virus was not reported in that sample. Reflex PCR results were evaluated separately and did not redefine the routine diagnostic ground truth used for cut-off optimization.

For each platform, 100,000 candidate cut-off sets were generated. AVITI candidate sets included minimum bleed ratio, minimum genome coverage, and minimum reads per million filtered reads, sampled across the following ranges: bleed ratio 0–10%, genome coverage 0–50%, and abundance 0–20 RPM. ONT candidate sets included minimum bleed ratio, minimum genome coverage, and minimum raw read count, sampled across the following ranges: bleed ratio 0–10%, genome coverage 0–50%, and abundance 0–100 reads.

Bleed ratio was calculated separately for each viral species and specimen type as:

*bleed ratio (%) = 100 × reads for a species in a given sample / maximum read count for that species in any sample of the same specimen type.*

Bleed-ratio filtering was applied only to high-abundance donor taxa, defined as taxa with sufficient signal to plausibly generate cross-sample bleed or index hopping. For AVITI, donor taxa were defined as species with a maximum read count ≥100,000 and maximum abundance ≥1,000 RPM within a specimen type. For ONT, donor taxa were defined as species with a maximum read count ≥10,000 and maximum abundance ≥1,000 RPM (reads per million mapped reads) within a specimen type. This filtering was to ensure only "high viral load" situations being included in such framework.

#### 1.2.2. Cut-off scoring and selection

Each potential cut-off set was applied to each row of detections. Detection sensitivity was calculated as TP/(TP + FN), where TP (true positive) represented expected sample-virus detections retained after filtering and FN (false negative) represented expected detections removed by filtering.

A non-expected sample–virus detection was defined as a sequencing signal for a virus that was not reported by routine diagnostic testing in that sample. Such a detection was considered a false positive when it remained positive after application of a candidate cut-off set. Candidate cut-off sets were first restricted to those producing no retained virus detections in the negative clinical samples, corresponding to 100% sample-level specificity in the optimization dataset.

The remaining candidate sets were ranked deterministically using a predefined hierarchy. Detection sensitivity was prioritized first, followed by the minimum of detection sensitivity and sample-level specificity, their average, sample-level specificity, and lower overall cut-off burden. The highest-ranked candidate set was selected as the operational cut-off for each platform.

To evaluate robustness, the top 100 ranked cut-off sets were retained and their clinical performance was recalculated. The distribution of these top-performing sets, the median top-100 parameter profile, and the final selected operational cut-offs are shown in **Supplementary Figure 1.**

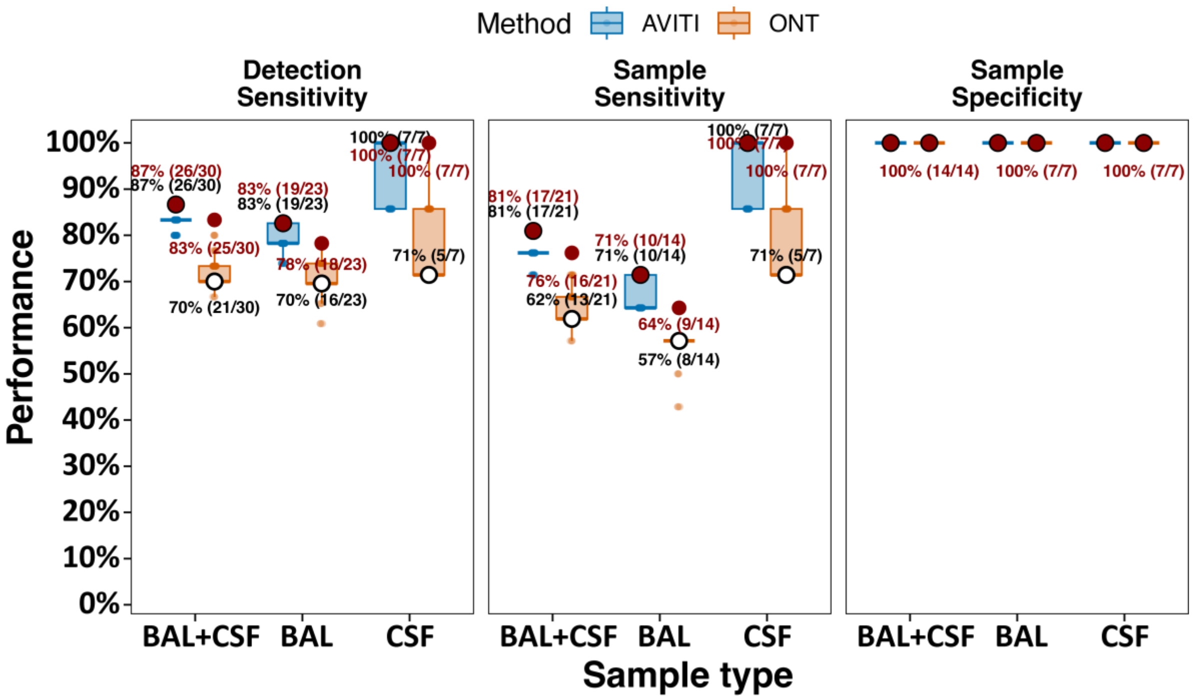


**Supplementary Figure 1.** Robustness of clinical performance across the top 100 LHS-derived cutoff sets. Routine clinical performance was recalculated for the top 100 feasible cutoff sets selected by the predefined LHS ranking procedure. Performance is shown for BAL and CSF combined, BAL only, and CSF only. Each small point represents the result of one cutoff set, and boxplots summarize the distribution across the top 100 sets. White circles indicate the performance obtained using the median parameter values of the top 100 cutoff sets. Red circles indicate the final selected operational cutoff set used for the main analyses. Labels show percentage and numerator/denominator.

#### 1.2.3. External validation of the operational cut-offs

The numerical operational cut-off values were transferred unchanged to the independent EQA datasets from 2024 and 2025. These panels were not used to select the operational cut-offs. Metagenomics performance was evaluated using various metrics as explained below.

**Detection sensitivity** was calculated across 23 expected sample–virus pairs spanning eight viral targets. Off-target removal was evaluated within the same eight-virus QCMD target panel. A raw off-target detection was defined as an observed sequencing signal for a panel virus in an EQA sample in which that virus was not expected. **Off-target removal** was calculated as the proportion of these observed raw off-target detections that no longer satisfied the operational cut-offs.

For the AVITI Human mastadenovirus E record corresponding to HadV-4, which was present at high abundance in a co-sequenced sample, the bleed-ratio denominator was set to the maximum read count observed in that sample rather than the maximum within the EQA panel.

After application of the selected LHS-derived cut-offs, detection-level sensitivity was 95.7% (22/23) for both AVITI and ONT. AVITI removed 53 of 56 observed raw off-target detections (94.6%), whereas ONT removed 29 of 30 (96.7%). Both platforms correctly classified the two negative EQA samples after filtering (2/2). The single expected target not retained by AVITI was HadV-4 in 2025 Sample 6, whereas the single expected target not retained by ONT was HSV-2 in 2024 Sample 6.

#### 1.2.4. Use of extraction negative controls for filtering

In addition to the sampled cut-off parameters explained above, a negative-control baseline filter was applied. For each viral species and specimen type, the maximum genome coverage observed in negative controls was used as a species-specific "background" threshold. A detection was retained only if its genome coverage was at least as high as this negative-control maximum.

**Supplementary Table 1.** Samples selected for testing with viral metagenomics. Blue indicates reflex PCR performed after review of the initial metagenomic results. Red indicates an initial routine-positive result that was not confirmed by subsequent reflex PCR and was therefore excluded from the reported clinical performance analysis. “Negative” indicates that no viral pathogen was reported during the original routine diagnostic work-up; subsequent reflex PCR findings did not alter this original classification.

| *ID* | *Sample type* | *Routine diagnostic status* | *Virus 1* | *Cq* | *Virus 2* | *Cq* | *Virus 3* | *Cq* | *Virus 4* | *Cq* | *could not be tested for* |
|---|---|---|---|---|---|---|---|---|---|---|---|
| *C1* | CSF | positive | VZV | 26.3 | | | | | | | |
| *C2* | CSF | positive | HSV-1 | 29.3 | EBV | 28 | | | | | |
| *C3* | CSF | positive | HHV-6 | 28.9 | | | | | | | |
| *C4* | CSF | positive | Polyoma | 2.87 log copies/ml | HHV-6 | 33 | | | | | |
| *C5* | CSF | positive | HSV-2 | 33.9 | | | | | | | |
| *C6* | CSF | positive | Echovirus 30 | 32.1 | | | | | | | |
| *C7* | CSF | positive | EBV | 2.01 log IU/ml | | | | | | | |
| *C8* | CSF | negative | - | 17.2 | | | | | | | |
| *C9* | CSF | negative | - | 18.6 | | | | | | | |
| *C10* | CSF | negative | - | | | | | | | | |
| *C11* | CSF | negative | - | | | | | | | | |
| *C12* | CSF | negative | - | | | | | | | | |
| *C13* | CSF | Suspected for CNS infection | - | | | | | | | | |
| *C14* | CSF | Suspected for CNS infection | - | | | | | | | | |
| *B1* | BAL | positive | Para-influenza type 2 | 34.9 | CMV | 29.8 | Entero/rhino | 32.4 | | | |

| | | | | | | | | | | | |
|---|---|---|---|---|---|---|---|---|---|---|---|
| *B2* | BAL | positive | hMPV | 34.3 | EBV | 32 | | | | | |
| *B3* | BAL | positive | RSV | 41.9 | hMPV | 36.2 | HSV-1 | 36.1 | EBV | 30.2 | |
| *B4* | BAL | positive | HSV-1 | 36.6 | EBV | 30.9 | HPyV6 | positive | | | HHV-7 & HHV-6 |
| *B5* | BAL | positive | Influenza A | 21.2 | Entero/rhino | 19 | EBV | 22.5 | HHV-7 | positive | |
| *B6* | BAL | negative | EBV | 39.4 | HHV-6 | 38.8 | HHV-7 | positive | | | |
| *B7* | BAL | positive | Adeno | 33 | SARS-CoV-2 | 37.6 | EBV | 25.2 | HHV-6 | 39.4 | |
| *B8* | BAL | negative | EBV | 35.6 | HHV-6 | 36 | | | | | |
| *B9* | BAL | negative | HHV-6 | 31.5 | HHV-7 | positive | | | | | |
| *B10* | BAL | negative | HHV-6 | 35.2 | | | | | | | |
| *B11* | BAL | negative | HHV-7 | positive | | | | | | | |
| *B12* | BAL | positive | Para-influenza type 3 | 22 | HSV-1 | 22.9 | EBV | 29.2 | HHV-7 | positive | |
| *B13* | BAL | positive | Para-influenza type 1 | 35.3 | Entero/rhino | 14 | EBV | 32.7 | HHV-7 | positive | |
| *B14* | BAL | positive | Adeno | 34.9 | | | | | | | |
| *B15* | BAL | positive | Corona OC43 | 26 | RSV | 19.4 | EBV | 28.9 | HHV-6 | 35.4 | |
| *B16* | BAL | positive | Adeno | 32.3 | HSV-1 | 36.3 | HHV-6 | 35.7 | HHV-7 | positive | |
| *B17* | BAL | positive | Influenza A | 29.3 | HSV-1 | 22 | EBV | 23.6 | HHV-6 | 37.6 | |
| *B18* | BAL | positive | Corona NL63 | 25.3 | CMV | 25.1 | HHV-7 | positive | | | EBV & HHV-6 |
| *B19* | BAL | negative | HHV-6 | 34.9 | HHV-7 | positive | | | | | |
| *B20* | BAL | negative | EBV | positive | HHV-7 | positive | | | | | |
| *B21* | BAL | positive | Entero D68 | 33.5 | RSV | 38 | HHV-7 | positive | | | EBV & HHV-6 |
