## Supplementary tables for "Short- and long-read hybrid-capture viral metagenomics reach high sensitivity and specificity with data-driven cut-offs compared with routine diagnostics in CSF and BAL specimens"

**Supplementary Table 1. EQA expected (target) and unexpected (off-target) viral reads per sample-virus.**

| | | | | | AVITI | | | | ONT | | | |
|---|---|---|---|---|---|---|---|---|---|---|---|---|
| **year** | sample | virus | Viral load (log10) | Target or off-target | Reads mapped to the virus | Depth | Genome coverage | Total quality filtered reads | Reads mapped to the virus | Depth | Genome coverage | Total mapped reads to viruses |
| **2024** | Sample1 | HIV-1 | 4 | Target | 6222 | 98.19 | 53.8 | 10116512 | 389 | 11.94 | 96.26 | 179748 |
| **2024** | Sample1 | MPXV | 4 | Target | 4110549 | 3267.45 | 98.81 | 10116512 | 126419 | 153.4 | 99.9 | 179748 |
| **2024** | Sample1 | CMV | | Off-target | | | | | 2 | 1 | 0.12 | 179748 |
| **2024** | Sample1 | EVD68 | | Off-target | 8 | 0.17 | 12.19 | 10116512 | | | | |
| **2024** | Sample1 | HSV-1 | | Off-target | 8 | 0.01 | 0.81 | 10116512 | | | | |
| **2024** | Sample1 | HSV-2 | | Off-target | 18 | 0.03 | 1.42 | 10116512 | 9 | 1 | 1.7 | 179748 |
| **2024** | Sample1 | Influenza A | | Off-target | 1 | 0.07 | 6.5 | 10116512 | 1 | 1 | 6.43 | 179748 |
| **2024** | Sample2 | EVD68 | 4.5 | Target | 46298 | 977.62 | 99.03 | 5011786 | 3152 | 104.65 | 97.93 | 161965 |
| **2024** | Sample2 | HSV-2 | 3.4 | Target | 779129 | 758.76 | 98.24 | 5011786 | 43106 | 78.02 | 92.44 | 161965 |
| **2024** | Sample2 | Influenza A | 5.7 | Target | 29564 | 407.47 | 79.65 | 5011786 | 619 | 91.46 | 97.51 | 161965 |
| **2024** | Sample2 | CMV | | Off-target | 14 | 0.01 | 0.92 | 5011786 | | | | |
| **2024** | Sample2 | HSV-1 | | Off-target | 2227 | 2.45 | 2.43 | 5011786 | | | | |
| **2024** | Sample2 | MPXV | | Off-target | 137 | 0.11 | 10.15 | 5011786 | 39 | 1.01 | 4.83 | 161965 |
| **2024** | Sample3 | CMV | 4 | Target | 116894 | 75.73 | 47.41 | 4522134 | 8002 | 25.34 | 32.77 | 144685 |
| **2024** | Sample3 | EVD68 | | Off-target | 14 | 0.29 | 24.47 | 4522134 | | | | |
| **2024** | Sample3 | HSV-1 | | Off-target | 1 | 0 | 0.13 | 4522134 | | | | |

| | | | | | | | | | | | | |
|---|---|---|---|---|---|---|---|---|---|---|---|---|
| **2024** | Sample3 | HSV-2 | | Off-target | 54 | 0.06 | 5.24 | 4522134 | | | | |
| **2024** | Sample3 | Influenza A | | Off-target | 7 | 0.1 | 6.23 | 4522134 | 1 | 1 | 5.17 | 144685 |
| **2024** | Sample3 | MPXV | | Off-target | 192 | 0.17 | 9.91 | 4522134 | 51 | 1.21 | 4.85 | 144685 |
| **2024** | Sample4 | EVD68 | 4.5 | Target | 40453 | 855.84 | 98.92 | 3824684 | 3793 | 132.62 | 95.4 | 138936 |
| **2024** | Sample4 | HSV-1 | 3.4 | Target | 83098 | 83.14 | 52.09 | 3824684 | 5681 | 24.75 | 38.37 | 138936 |
| **2024** | Sample4 | MPXV | 3 | Target | 67210 | 54.47 | 54.3 | 3824684 | 8113 | 23.06 | 41.04 | 138936 |
| **2024** | Sample4 | CMV | | Off-target | 1 | 0 | 0.06 | 3824684 | | | | |
| **2024** | Sample4 | HSV-2 | | Off-target | | | | | 53 | 3.21 | 1.58 | 138936 |
| **2024** | Sample4 | Influenza A | | Off-target | 625 | 8.59 | 3.57 | 3824684 | 44 | 21.85 | 21.48 | 138936 |
| **2024** | Sample5 | CMV | | Off-target | 3 | 0.02 | 0.06 | 3840310 | 2 | 1 | 0.25 | 140847 |
| **2024** | Sample5 | EVD68 | | Off-target | 119 | 2.48 | 6.18 | 3840310 | 16 | 6.83 | 4.8 | 140847 |
| **2024** | Sample5 | HSV-1 | | Off-target | 2 | 0 | 0.21 | 3840310 | 2 | 1 | 0.22 | 140847 |
| **2024** | Sample5 | HSV-2 | | Off-target | 8 | 0.01 | 0.96 | 3840310 | 29 | 2.27 | 1.4 | 140847 |
| **2024** | Sample5 | Influenza A | | Off-target | | | | | 1 | 1 | 8.18 | 140847 |
| **2024** | Sample5 | MPXV | | Off-target | 92 | 0.07 | 7.01 | 3840310 | 55 | 1.03 | 6.49 | 140847 |
| **2024** | Sample6 | EVD68 | 4.5 | Target | 57071 | 1189.22 | 98.72 | 4223834 | 5058 | 178.44 | 97.05 | 147531 |
| **2024** | Sample6 | HSV-1 | 1.4 | Target | 10987 | 10.64 | 7.05 | 4223834 | 799 | 16.77 | 9.12 | 147531 |
| **2024** | Sample6 | HSV-2 | 1.4 | Target | 10286 | 10.04 | 6.59 | 4223834 | 742 | 17.08 | 7.9 | 147531 |
| **2024** | Sample6 | CMV | | Off-target | 3 | 0 | 0.2 | 4223834 | 2 | 1 | 0.26 | 147531 |
| **2024** | Sample6 | Influenza A | | Off-target | | | | | 1 | 1 | 6.81 | 147531 |
| **2024** | Sample6 | MPXV | | Off-target | 250 | 0.21 | 17.53 | 4223834 | 51 | 1.05 | 6.22 | 147531 |
| **2025** | Sample1 | EVD68 | 4.5 | Target | 157565 | 3212.38 | 99.8 | 15476661 | 1170 | 43.93 | 85.2 | 48044 |
| **2025** | Sample1 | HSV-2 | 3.4 | Target | 2408394 | 2324.87 | 98.75 | 15476661 | 12675 | 24.55 | 75.17 | 48044 |
| **2025** | Sample1 | Influenza A | 5.7 | Target | 54972 | 755.01 | 57.4 | 15476661 | 111 | 17.23 | 72.86 | 48044 |
| **2025** | Sample1 | CMV | | Off-target | 110 | 0.07 | 5.07 | 15476661 | 2 | 1 | 0.15 | 48044 |
| **2025** | Sample1 | HIV-1 | | Off-target | 12 | 0.19 | 9.82 | 15476661 | | | | |

| | | | | | | | | | | | | |
|---|---|---|---|---|---|---|---|---|---|---|---|---|
| **2025** | Sample1 | HSV-1 | | Off-target | 13847 | 14.42 | 6.15 | 15476661 | | | | |
| **2025** | Sample1 | HadV-4 | | Off-target | 874 | 3.48 | 46.12 | 15476661 | | | | |
| **2025** | Sample1 | MPXV | | Off-target | 7016 | 5.35 | 93.28 | 15476661 | 24 | 1.01 | 3.06 | 48044 |
| **2025** | Sample2 | CMV | | Off-target | 55 | 0.03 | 3.04 | 8798185 | | | | |
| **2025** | Sample2 | EVD68 | | Off-target | 37 | 0.73 | 39.23 | 8798185 | | | | |
| **2025** | Sample2 | HIV-1 | | Off-target | 7 | 0.1 | 7.04 | 8798185 | | | | |
| **2025** | Sample2 | HSV-2 | | Off-target | 145 | 0.17 | 11.5 | 8798185 | | | | |
| **2025** | Sample2 | HadV-4 | | Off-target | 274 | 1.07 | 35.52 | 8798185 | | | | |
| **2025** | Sample2 | Influenza A | | Off-target | 7 | 0.09 | 7.58 | 8798185 | | | | |
| **2025** | Sample2 | MPXV | | Off-target | 2415 | 1.84 | 74.82 | 8798185 | 5 | 1 | 0.44 | 23643 |
| **2025** | Sample3 | HIV-1 | 4 | Target | 30978 | 456.77 | 61.71 | 25509167 | 862 | 36.47 | 57.93 | 165233 |
| **2025** | Sample3 | MPXV | 4 | Target | 10396358 | 8128.82 | 98.88 | 25509167 | 125660 | 141.67 | 99.93 | 165233 |
| **2025** | Sample3 | CMV | | Off-target | 71 | 0.05 | 3.51 | 25509167 | 1 | 1 | 0.12 | 165233 |
| **2025** | Sample3 | EVD68 | | Off-target | 62 | 1.24 | 51.09 | 25509167 | | | | |
| **2025** | Sample3 | HSV-1 | | Off-target | 3 | 0.01 | 0.36 | 25509167 | | | | |
| **2025** | Sample3 | HSV-2 | | Off-target | 271 | 0.4 | 18.41 | 25509167 | 18 | 2.14 | 0.89 | 165233 |
| **2025** | Sample3 | HadV-4 | | Off-target | 570 | 2.24 | 44.4 | 25509167 | | | | |
| **2025** | Sample3 | Influenza A | | Off-target | 10 | 0.15 | 12.69 | 25509167 | 1 | 1 | 17.86 | 165233 |
| **2025** | Sample4 | EVD68 | 4.5 | Target | 106810 | 2198.69 | 99.77 | 8868193 | 783 | 37.19 | 63.48 | 22305 |
| **2025** | Sample4 | HSV-1 | 1.4 | Target | 21723 | 21.67 | 5.26 | 8868193 | 110 | 3.72 | 5.03 | 22305 |
| **2025** | Sample4 | HSV-2 | 1.4 | Target | 16821 | 16.76 | 19.2 | 8868193 | 20592 | 209.5 | 4.3 | 22305 |
| **2025** | Sample4 | Influenza A | 4.7 | Target | 10313 | 144.83 | 12.01 | 8868193 | 90 | 50.24 | 13.38 | 22305 |
| **2025** | Sample4 | CMV | | Off-target | 56 | 0.03 | 2.78 | 8868193 | | | | |
| **2025** | Sample4 | HIV-1 | | Off-target | 6 | 0.09 | 7.83 | 8868193 | | | | |
| **2025** | Sample4 | HadV-4 | | Off-target | 377 | 1.49 | 39.66 | 8868193 | | | | |
| **2025** | Sample4 | MPXV | | Off-target | 3015 | 2.3 | 80.81 | 8868193 | 1 | 1 | 0.1 | 22305 |

| | | | | | | | | | | | | |
|---|---|---|---|---|---|---|---|---|---|---|---|---|
| **2025** | Sample5 | CMV | 4 | Target | 402611 | 255.93 | 43.31 | 12898543 | 34318 | 29.52 | 45.65 | 35666 |
| **2025** | Sample5 | EVD68 | | Off-target | 55 | 1.09 | 51.66 | 12898543 | | | | |
| **2025** | Sample5 | HIV-1 | | Off-target | 8 | 0.13 | 3.97 | 12898543 | | | | |
| **2025** | Sample5 | HSV-1 | | Off-target | 4 | 0 | 0.46 | 12898543 | | | | |
| **2025** | Sample5 | HSV-2 | | Off-target | 198 | 0.26 | 14.81 | 12898543 | 12 | 3.54 | 0.32 | 35666 |
| **2025** | Sample5 | HadV-4 | | Off-target | 459 | 1.82 | 41.1 | 12898543 | | | | |
| **2025** | Sample5 | Influenza A | | Off-target | 17 | 0.22 | 14.28 | 12898543 | 1 | 1 | 13.15 | 35666 |
| **2025** | Sample5 | MPXV | | Off-target | 3726 | 2.85 | 85.96 | 12898543 | 5 | 1 | 0.61 | 35666 |
| **2025** | Sample6* | HadV-4 | 3.3 | Target | 2679 | 10.89 | 47.99 | 39569079 | 7 | 1 | 4.36 | 151473 |
| **2025** | Sample6* | CMV | | Off-target | 27 | 0.02 | 1.57 | 39569079 | 3 | 1.34 | 0.2 | 151473 |
| **2025** | Sample6* | EVD68 | | Off-target | 18 | 0.36 | 24.93 | 39569079 | | | | |
| **2025** | Sample6* | HIV-1 | | Off-target | 5 | 0.08 | 4.99 | 39569079 | | | | |
| **2025** | Sample6* | HSV-2 | | Off-target | 70 | 0.08 | 5.68 | 39569079 | 8 | 1.12 | 1.12 | 151473 |
| **2025** | Sample6* | Influenza A | | Off-target | 1 | 0.02 | 1.82 | 39569079 | | | | |
| **2025** | Sample6* | MPXV | | Off-target | 1338 | 1.01 | 56.86 | 39569079 | 97 | 1.1 | 11.62 | 151473 |

**Supplementary table 2. Virus species that were not presented in the manuscript and the reason of not including such virus species.**

| Species | Host category | Filtering reason |
|---|---|---|
| Duck aviadenovirus B | Animal - environmental | Potential mismapping / low level multiple sample presence |
| Fowl aviadenovirus E | Animal - environmental | Potential mismapping / low level multiple sample presence |
| Pigeon aviadenovirus A | Animal - environmental | Potential mismapping / low level multiple sample presence |
| Torque teno virus 1 | Human | Not directly related to disease |
| Torque teno virus 10 | Human | Not directly related to disease |
| Torque teno virus 11 | Human | Not directly related to disease |
| Torque teno virus 12 | Human | Not directly related to disease |
| Torque teno virus 13 | Human | Not directly related to disease |
| Torque teno virus 15 | Human | Not directly related to disease |
| Torque teno virus 16 | Human | Not directly related to disease |
| Torque teno virus 17 | Human | Not directly related to disease |
| Torque teno virus 18 | Human | Not directly related to disease |
| Torque teno virus 19 | Human | Not directly related to disease |
| Torque teno virus 20 | Human | Not directly related to disease |
| Torque teno virus 21 | Human | Not directly related to disease |
| Torque teno virus 22 | Human | Not directly related to disease |
| Torque teno virus 23 | Human | Not directly related to disease |
| Torque teno virus 24 | Human | Not directly related to disease |
| Torque teno virus 27 | Human | Not directly related to disease |
| Torque teno virus 28 | Human | Not directly related to disease |
| Torque teno virus 29 | Human | Not directly related to disease |
| Torque teno virus 3 | Human | Not directly related to disease |

| | | |
|---|---|---|
| Torque teno virus 5 | Human | Not directly related to disease |
| Torque teno virus 6 | Human | Not directly related to disease |
| Torque teno virus 7 | Human | Not directly related to disease |
| Torque teno virus 8 | Human | Not directly related to disease |
| Torque teno virus 9 | Human | Not directly related to disease |
| TTV-like mini virus | Human | Not directly related to disease |
| Torque teno mini virus 1 | Human | Not directly related to disease |
| Torque teno mini virus 12 | Human | Not directly related to disease |
| Torque teno mini virus 27 | Human | Not directly related to disease |
| Torque teno mini virus 32 | Human | Not directly related to disease |
| Torque teno mini virus 7 | Human | Not directly related to disease |
| Torque teno mini virus 8 | Human | Not directly related to disease |
| Torque teno mini virus 9 | Human | Not directly related to disease |
| Torque teno midi virus 1 | Human | Not directly related to disease |
| Torque teno midi virus 10 | Human | Not directly related to disease |
| Torque teno midi virus 12 | Human | Not directly related to disease |
| Torque teno midi virus 13 | Human | Not directly related to disease |
| Torque teno midi virus 14 | Human | Not directly related to disease |
| Torque teno midi virus 2 | Human | Not directly related to disease |
| Torque teno midi virus 3 | Human | Not directly related to disease |
| Torque teno midi virus 4 | Human | Not directly related to disease |
| Torque teno midi virus 5 | Human | Not directly related to disease |
| Torque teno midi virus 6 | Human | Not directly related to disease |
| Torque teno midi virus 8 | Human | Not directly related to disease |
| Torque teno midi virus 9 | Human | Not directly related to disease |
| Small anellovirus | Human | Not directly related to disease |
| Torque teno virus | Human | Not directly related to disease |

| | | |
|---|---|---|
| Sapporo virus | Human | Another sample with high viral load was sequenced in the same run |
| Giant panda circovirus 1 | Animal - environmental | Not directly related to disease |
| Human associated cyclovirus 10 | Human | Not directly related to disease |
| Circovirus-like genome DCCV-10 | Contaminant | Known contaminant |
| Circovirus-like genome DCCV-13 | Contaminant | Known contaminant |
| Circovirus-like genome DCCV-4 | Contaminant | Known contaminant |
| Circovirus-like genome DCCV-7 | Contaminant | Known contaminant |
| Israeli acute paralysis virus | Animal - environmental | Insect-infecting virus, not clinically relevant. |
| Yellow fever virus | Human | Another sample with high viral load was sequenced in the same run |
| Pegivirus C | Human | Not directly related to disease |
| Pestivirus A | Animal - environmental | Not directly related to disease |
| Pestivirus G | Animal - environmental | Not directly related to disease |
| Paslahepevirus balayani | Human - animal | Another sample with high viral load was sequenced in the same run |
| Rocahepevirus ratti | Human - animal | Potential mismapping |
| Aotine betaherpesvirus 1 | Human -animal | Potential mismapping / low level multiple sample presence |
| Cercopithecine betaherpesvirus 5 | Human -animal | Potential mismapping / low level multiple sample presence |
| Panine betaherpesvirus 2 | Human -animal | Potential mismapping / low level multiple sample presence |
| Papiine betaherpesvirus 3 | Human -animal | Potential mismapping / low level multiple sample presence |
| Macacine gammaherpesvirus 10 | Human -animal | Potential mismapping / low level multiple sample presence |

| | | |
|---|---|---|
| Macacine gammaherpesvirus 4 | Human -animal | Potential mismapping / low level multiple sample presence |
| Ateline gammaherpesvirus 3 | Human -animal | Potential mismapping / low level multiple sample presence |
| Bovine gammaherpesvirus 4 | Human -animal | Potential mismapping / low level multiple sample presence |
| Common bottlenose dolphin gammaherpesvirus 1 strain Sarasota | Human -animal | Potential mismapping / low level multiple sample presence |
| Human gammaherpesvirus 8 | Human -animal | Potential mismapping / low level multiple sample presence |
| Macacine gammaherpesvirus 8 | Human -animal | Potential mismapping / low level multiple sample presence |
| Macacine betaherpesvirus 9 | Human -animal | Potential mismapping / low level multiple sample presence |
| Ateline alphaherpesvirus 1 | Human -animal | Potential mismapping / low level multiple sample presence |
| Bovine alphaherpesvirus 2 | Human -animal | Potential mismapping / low level multiple sample presence |
| Cercopithecine alphaherpesvirus 2 | Human -animal | Potential mismapping / low level multiple sample presence |
| Macacine alphaherpesvirus 1 | Human -animal | Potential mismapping / low level multiple sample presence |
| Macropodid alphaherpesvirus 1 | Human -animal | Potential mismapping / low level multiple sample presence |
| Macropodid alphaherpesvirus 2 | Human -animal | Potential mismapping / low level multiple sample presence |
| Panine alphaherpesvirus 3 | Human -animal | Potential mismapping / low level multiple sample presence |
| Papiine alphaherpesvirus 2 | Human -animal | Potential mismapping / low level multiple sample presence |

| | | |
|---|---|---|
| Saimiriine alphaherpesvirus 1 | Human -animal | Potential mismapping / low level multiple sample presence |
| Bovine alphaherpesvirus 1 | Human -animal | Potential mismapping / low level multiple sample presence |
| Bovine alphaherpesvirus 5 | Human -animal | Potential mismapping / low level multiple sample presence |
| Bubaline alphaherpesvirus 1 | Human -animal | Potential mismapping / low level multiple sample presence |
| Caprine alphaherpesvirus 1 | Human -animal | Potential mismapping / low level multiple sample presence |
| Cervid alphaherpesvirus 1 | Human -animal | Potential mismapping / low level multiple sample presence |
| Cervid alphaherpesvirus 2 | Human -animal | Potential mismapping / low level multiple sample presence |
| Equid alphaherpesvirus 1 | Human -animal | Potential mismapping / low level multiple sample presence |
| Equid alphaherpesvirus 3 | Human -animal | Potential mismapping / low level multiple sample presence |
| Equid alphaherpesvirus 4 | Human -animal | Potential mismapping / low level multiple sample presence |
| Monodontid alphaherpesvirus 1 | Human -animal | Potential mismapping / low level multiple sample presence |
| Suid alphaherpesvirus 1 | Human -animal | Potential mismapping / low level multiple sample presence |
| Varroa destructor virus 1 | Animal - environmental | Insect-infecting virus, not clinically relevant. |
| Alphapapillomavirus 10 | Human | Not directly related to disease in BAL-CSF samples |
| Alphapapillomavirus 14 | Human | Not directly related to disease in BAL-CSF samples |

| | | |
|---|---|---|
| Betapapillomavirus 1 | Human | Not directly related to disease in BAL-CSF samples |
| Betapapillomavirus 2 | Human | Not directly related to disease in BAL-CSF samples |
| Betapapillomavirus 3 | Human | Not directly related to disease in BAL-CSF samples |
| Betapapillomavirus 5 | Human | Not directly related to disease in BAL-CSF samples |
| Dyoomikronpapillomavirus 1 | Human | Not directly related to disease in BAL-CSF samples |
| Gammapapillomavirus 10 | Human | Not directly related to disease in BAL-CSF samples |
| Gammapapillomavirus 12 | Human | Not directly related to disease in BAL-CSF samples |
| Gammapapillomavirus 15 | Human | Not directly related to disease in BAL-CSF samples |
| Gammapapillomavirus 22 | Human | Not directly related to disease in BAL-CSF samples |
| Gammapapillomavirus 24 | Human | Not directly related to disease in BAL-CSF samples |
| Gammapapillomavirus 5 | Human | Not directly related to disease in BAL-CSF samples |
| Gammapapillomavirus 8 | Human | Not directly related to disease in BAL-CSF samples |
| Omegapapillomavirus 1 | Human | Not directly related to disease in BAL-CSF samples |
| Adeno-associated dependoparvovirus A | Human | Not directly related to disease in BAL-CSF samples |
| Primate protoparvovirus 3 | Human | Not directly related to disease in BAL-CSF samples (skin associated) |

| | | |
|---|---|---|
| Parus major densovirus | Animal - environmental | Known contaminant/potential mismapping |
| Parvovirus NIH-CQV | Contaminant | Known contaminant |
| Tete orthobunyavirus | Animal - environmental | Potential mismapping / low level multiple sample presence |
| Cardiovirus A | Human | Potential mismapping from other high viral load enteroviruses / low level multiple sample presence |
| Enterovirus A | Human | Potential mismapping from other high viral load enteroviruses / low level multiple sample presence |
| Enterovirus C | Human | Potential mismapping from other high viral load enteroviruses / low level multiple sample presence |
| Human echovirus AMS721 | Human | Potential mismapping from other high viral load enteroviruses / low level multiple sample presence |
| Rhinovirus A | Human | Potential mismapping from other high viral load enteroviruses / low level multiple sample presence |
| Alphapolyomavirus quintihominis | Human | Not directly related to disease in BAL-CSF samples |
| Betapolyomavirus macacae | Animal | Potential mismapping / low level multiple sample presence |
| Deltapolyomavirus decihominis | Human | Not directly related to disease in BAL-CSF samples |
| Deltapolyomavirus undecihominis | Human | Not directly related to disease in BAL-CSF samples |
| Molluscum contagiosum virus | Human | Not directly related to disease in BAL-CSF samples |

| | | |
|---|---|---|
| Cowpox virus | Animal | Potential mismapping / low level multiple sample presence |
| Pseudocowpox virus | Animal | Potential mismapping / low level multiple sample presence |
| Avian leukosis virus | Animal-potentially endogenous viral element | Potential mismapping / low level multiple sample presence |
| Avian myeloblastosis virus | Animal-potentially endogenous viral element | Potential mismapping / low level multiple sample presence |
| Avian myelocytomatosis virus 29 | Animal-potentially endogenous viral element | Potential mismapping / low level multiple sample presence |
| Rous sarcoma virus | Animal-potentially endogenous viral element | Potential mismapping / low level multiple sample presence |
| Jaagsiekte sheep retrovirus | Animal-potentially endogenous viral element | Potential mismapping / low level multiple sample presence |
| Mouse mammary tumor virus | Animal-potentially endogenous viral element | Potential mismapping / low level multiple sample presence |
| Ovine enzootic nasal tumor virus | Animal-potentially endogenous viral element | Potential mismapping / low level multiple sample presence |
| Abelson murine leukemia virus | Animal-potentially endogenous viral element | Potential mismapping / low level multiple sample presence |
| Hardy-Zuckerman feline sarcoma virus | Animal-potentially endogenous viral element | Potential mismapping / low level multiple sample presence |
| Harvey murine sarcoma virus | Animal-potentially endogenous viral element | Potential mismapping / low level multiple sample presence |
| Kirsten murine sarcoma virus | Animal-potentially endogenous viral element | Potential mismapping / low level multiple sample presence |
| Murine leukemia virus | Animal-potentially endogenous viral element | Potential mismapping / low level multiple sample presence |
| Murine leukemia-related retroviruses | Animal-potentially endogenous viral element | Potential mismapping / low level multiple sample presence |

| | | |
|---|---|---|
| Murine osteosarcoma virus | Animal-potentially endogenous viral element | Potential mismapping / low level multiple sample presence |
| Mus musculus mobilized endogenous polytropic provirus | Animal-potentially endogenous viral element | Potential mismapping / low level multiple sample presence |
| Porcine type-C oncovirus | Animal-potentially endogenous viral element | Potential mismapping / low level multiple sample presence |
| Snyder-Theilen feline sarcoma virus | Animal-potentially endogenous viral element | Potential mismapping / low level multiple sample presence |
| Spleen focus-forming virus | Animal-potentially endogenous viral element | Potential mismapping / low level multiple sample presence |
| Woolly monkey sarcoma virus | Animal-potentially endogenous viral element | Potential mismapping / low level multiple sample presence |
| Human immunodeficiency virus 2 | Animal-potentially endogenous viral element | Potential mismapping / low level multiple sample presence |
| Avian endogenous retrovirus EAV-HP | Animal-potentially endogenous viral element | Potential mismapping / low level multiple sample presence |
| Human endogenous retrovirus K | Human-potentially endogenous viral element | Potential mismapping / low level multiple sample presence |
| Tomato torrado virus | Plant related - environment | Environmental |
| Rotavirus A | Human | Low level presence in multiple samples , known lab contaminant |
| Ball python nidovirus 1 | Animal- environmental | Potential mismapping / low level multiple sample presence |
| Pregotovirus heba | Animal- environmental | Potential mismapping / low level multiple sample presence |
